# Advanced HIV Disease Burden in Selected Health Facilities in Mozambique, October–December 2023

**DOI:** 10.1101/2024.11.26.24317679

**Authors:** Alexandre Nguimfack, Eudoxia Filipe, Dercio Filimao, José Mizela, Erica Bila, Mercia Matsinhe, Maria Inês Tomo de Deus, Irenio Gaspar, Maria Ruano, Luis Armando, Orrin Tiberi

## Abstract

**Introduction:** Advanced HIV Disease (AHD) is a remaining hurdle in the care and treatment cascade in Mozambique and has the potential to delay Mozambique’s progress towards the UNAIDS 95-95-95 goals for 2025. An improved understanding of the prevalence and demographics of AHD is critical to understanding this important condition.

**Methods:** Data from October to December 2023 were reviewed from the 79 health facilities implementing the AHD diagnosis and treatment package at the national level.

**Results:** Overall, 36% (7.811/21.452) of people living with HIV (PLHIV) newly initiated or reinitiated on treatment had a CD4 test, of which 1.986 were diagnosed with AHD. Most diagnosed cases were in the newly initiated PLHIV (1.814) versus reinitiated (179).

**Conclusion:** Low CD4 testing uptake means that AHD remains a serious challenge in Mozambique’s path towards the elimination of HIV as a public health threat.

## Introduction

Mozambique has a high-burden of HIV, with a prevalence of 12.5% in 2021 and an estimated 2.4 million people living with HIV (PLHIV) in 2023.^1,2^ Over the past 20 years, scale-up of antiretroviral therapy (ART) has improved health outcomes and mortality rates among PLHIV and contributed to declining HIV-related deaths and new infections.^2,3^ Despite ART expansion, the adequate and timely identification and treatment of advanced HIV disease (AHD) remains a challenge. The World Health Organization (WHO) defines AHD as the Cluster of Differentiation 4 (CD4) cell count <200 cells/mm3 of blood or through coinfection diagnosis, or disease staging, in adults and adolescents. Previous studies found a prevalence of AHD in Mozambique and neighboring South Africa of 14.2% and 43.45%, respectively.^4,5^

In March 2022, the Ministry of Health began AHD diagnosis and treatment programming at selected sites in Mozambique by offering AHD screening, testing, and treatment. We analyzed programmatic data to understand the burden of AHD among PLHIV aged ≥5 years from October to December 2023.

## Methods

People were classified as being newly initiated on ART if they had HIV and enrolled on ART for the first time during the reporting period. People were classified as having reinitiated ART if they had HIV and had experienced an interruption in treatment for more than 60 days since the last expected drug pick-up, restarted ART during October–December 2023, and remained on ART by the end of the study period (December 20, 2023). The total number initiated on ART is the sum of people in these two treatment categories. CD4 coverage was calculated as the number of PLHIV who had a CD4 test among those who were eligible, according to their treatment status. Eligibility for CD4 testing includes any PLHIV six months after beginning treatment for the first time or upon reinitiating treatment. For this analysis, AHD was defined as PLHIV with a CD4 conducted during October–December 2023 with <200 cells/mm3 at the time of testing. Data were analyzed by sex, age group (i.e., 5–15, >15 years), patient category (newly-initiated, reinitiated), and CD4 results.^1^

Data were analyzed from the Electronic Patient Tracking System (EPTS) for 79 national AHD sites from October to December 2023. Any recipient of care who was less than 5 years old by December 20, 2023 was excluded from the analysis.

## Results

During October–December 2023, 21.452 PLHIV were eligible for CD4 screening at the 79 AHD sites. Of that sample, 12.156 (57%) were newly initiated, and 9.296 (43%) were reinitiated. Of the PLHIV newly initiated on ART, 94% (11.372) were over the age of 15, 38% (4.631) were men, and the mean age was 31 years (range = 7–67 years). Among the PLHIV reinitiated on ART, 39% (3.598) were men, 97% (9.055) were over the age of 15, and the mean age was 26 years (range = 7–67 years).

A total of 7,811 PLHIV were screened for AHD with a CD4 test during October–December 2023, or approximately 36% of the total number of people eligible. Among them, 90% (7,002) were newly initiated on ART, and 10% (809) were reinitiated on ART. CD4 coverage was much higher among newly initiated PLHIV (57.6% [7,002/12,156]) than among reinitiated PLHIV (9% [809/9,296]). CD4 coverage was slightly higher among men than women (37% vs. 36%) and higher in the 5-15 age group (43% [436/1,025]) than among those >15 years (36% [7,375/20,427]).

**Table 1:** CD4 value of newly initiated and reinitiating patients on ART in advanced HIV disease sites, Mozambique, October-December 2023.

| Indicators |  | Female | Male | 5-15 years | 15+ years | Total |
| --- | --- | --- | --- | --- | --- | --- |
| Reinitiated on ART | Reinitiated on ART | 5.698 | 3.598 | 241 | 9.055 | **9.296** |
|  | With CD4 Results | 502 | 307 | 32 | 777 | **809** |
|  | With CD4 Results <200 | 83 | 89 | 3 | 169 | **172** |
|  | % Reinitiated with CD4 Results | 9% | 9% | 13% | 9% | **9%** |
|  | **% of CD4 Results <200** | **17%** | **29%** | **9%** | **22%** | **21%** |
| Newly-Initiated on ART | Newly Initiated on ART | 7.525 | 4.631 | 784 | 11.372 | **12.156** |
|  | With CD4 Results | 4,252 | 2,750 | 404 | 6,598 | **7.002** |
|  | With CD4 Results <200 | 926 | 888 | 65 | 1,749 | **1.814** |
|  | % Newly Initiated with CD4 Results | 57% | 59% | 52% | 58% | **58%** |
|  | **% of CD4 Results <200** | **22%** | **32%** | **16%** | **27%** | **26%** |
| Total Initiated<br>(Reinitiated + Newly) | Total Initiated on ART | 13.223 | 8.229 | 1.025 | 20.427 | **21.452** |
|  | With CD4 Results | 4,754 | 3,057 | 436 | 7,375 | **7.811** |
|  | With CD4 Results <200 | 1,009 | 977 | 68 | 1,918 | **1.986** |
|  | % Total Initiated with CD4 Results | 36% | 37% | 43% | 36% | **36%** |
|  | **% of CD4 Results <200** | **21%** | **32%** | **16%** | **26%** | **25%** |

ART: Antiretroviral

A total of 1,986 of the 21.452 PLHIV were diagnosed with AHD during this reporting period; of those, 91.3% (1,814) were newly initiated, and 8.7% (172) were reinitiated. Men were 49% of the total (977/1,986), and the mean age was 34 years (range = 7–67 years). Among those with CD4 test results, 26% (1.814/7,002) of newly initiated PLHIV were diagnosed with AHD, compared to 21% (172/809) of reinitiated PLHIV. In the sample, a higher percentage of men were diagnosed with AHD (32% [977/3,057]) than women (21% [1,009/4,754]). The proportion with AHD was also higher among those aged >15 years (26% [1,918/7,375]) compared to children aged 5–15 years (16% [68/436]).

## Discussion

AHD was more common in men, supporting the increased focus on the engagement of men and the adoption of male health service use and ART retention strategies. Nevertheless, AHD was high among women; more than one out of five women screened in the sample had AHD. Data also showed low uptake within the 79 AHD sites; despite the availability of AHD clinical care, only approximately one-third of eligible PLHIV were tested with CD4, which is the entry point for AHD diagnosis and treatment. Low rates of CD4 testing, especially among reinitiated PLHIV, present a challenge for fully understanding the burden of AHD and ensuring appropriate clinical care. Expanding CD4 testing uptake and improving CD4 eligibility monitoring could contribute to improving the diagnosis of AHD and reduce HIV-related morbidity and mortality.

Our analysis is limited geographically. The number of AHD sites only represents 4.5% of the total sites implementing ART in Mozambique. In addition, we do not have outcome data for the PLHIV diagnosed with AHD to better understand the entire AHD management cascade.

## Conclusion

Despite overall progress in the national HIV response, AHD presents a serious challenge in Mozambique with a high proportion of newly initiated and reinitiated PLHIV diagnosed with AHD. Patients with advanced HIV disease are more vulnerable to poorer health outcomes, including death, and present a greater strain on healthcare resources.

## Data Availability

All data produced are available from PEPFAR Monitoring, Evaluation, and Reporting program data.

https://data.pepfar.gov/datasets#PDD

## Acknowledgements

We would like to acknowledge the support of PEPFAR Mozambique and the Mozambican Ministry of Health – National Public Health Directorate in the conception, elaboration, and approval of this short report.

## Funding Acknowledgement

This short-report has been supported by the President’s Emergency Plan for AIDS Relief (PEPFAR), through the Centers for Disease Control and Prevention (CDC). The findings and conclusions in this short-report are those of the authors and do not necessarily represent the official position of the funding agencies.

## Footnotes

1 This activity was reviewed by CDC, deemed not research, and was conducted consistent with applicable federal law and CDC policy. See e.g., 45 C.F.R. part 46.102(I)(2), 21 C.F.R. part 56; 42 U.S.C. 241(d); 5 U.S.C. 552a; 44 U.S.C. 3501 et seq.

